# Modelling the impact of lockdown easing measures on cumulative COVID-19 cases and deaths in England

**DOI:** 10.1101/2020.06.21.20136853

**Authors:** H Ziauddeen, N Subramaniam, D Gurdasani

**Affiliations:** Dept. of Psychiatry, University of Cambridge, Cambridge UK; Wellcome Trust-MRC Institute of Metabolic Science, University of Cambridge, Cambridge UK; Cambridgeshire & Peterborough Foundation Trust, Cambridge UK; William Harvey Research Institute, Barts and The London School of Medicine and Dentistry, Queen Mary University of London, London UK

**Author notes:** **Corresponding author:** Dr Deepti Gurdasani, William Harvey Research Institute, Barts and The London School of Medicine and Dentistry, Queen Mary University of London, London UK.

## Abstract

**Background:** As countries begin to ease the lockdown measures instituted to control the COVID-19 pandemic, there is a risk of a resurgence of the pandemic, and early reports of this are already emerging from some countries. Unlike many other countries, the UK started easing lockdown in England when levels of community transmission were still high, and this could have a major impact on case numbers and deaths. However thus far, the likely impacts of easing restrictions at this point in the pandemic have not been quantified. Using a Bayesian model, we assessed the potential impacts of successive lockdown easing measures in England, focussing on scenarios where the reproductive number (*R*) remains ≤1 in line with the UK government’s stated aim.

**Methods:** We developed a Bayesian model to infer incident cases and *R* in England, from incident death data from the Office of National Statistics. We then used this to forecast excess cases and deaths in multiple plausible scenarios in which *R* increases at one or more time points, compared to a baseline scenario where *R* remains unchanged by the easing of lockdown.

**Findings:** The model inferred an *R* of 0.752 on the 13^th^ May when England first started easing lockdown. In the most conservative scenario where *R* increases to 0.80 as lockdown was eased further on 1^st^ June and then remained constant, the model predicts an excess 257 (95% 108-492) deaths and 26,447 (95% CI 11,105-50,549) cumulative cases over 90 days. In the scenario with maximal increases in *R* (but staying ≤1) with successive easing of lockdown, the model predicts 3,174 (95% 1,334-6,060) excess cumulative deaths and 421,310 (95% 177,012-804,811) excess cases.

**Results:** When levels of transmission are high, even small changes in *R* with easing of lockdown can have significant impacts on expected cases and deaths, even if *R* remains ≤1. This will have a major impact on population health, tracing systems and health care services in England.. Following an elimination strategy rather than one of maintenance of *R* below 1 would substantially mitigate the impact of the COVID-19 epidemic within England. This study provides urgently needed information for developing public health policy for the next stages of the pandemic.

## Introduction

As countries around the world negotiate the first wave of the COVID-19 pandemic, governments have had to make critical decisions about when and how they ease the lockdown measures that were instituted to control the pandemic. Given the risks of a resurgence of the pandemic and the consequent implications, these decisions need to be informed by best available scientific evidence available at the time.

Different countries have eased lockdown in different ways, and at different points in their epidemic trajectory.^1^ The UK imposed lockdown relatively late in its epidemic trajectory and began easing lockdown relatively early, when community transmission levels (incident cases) were still high.^2^ By contrast, Germany, Denmark, Italy and Spain started easing lockdown when incident cases and deaths were at much lower levels. Despite mitigating strategies such as test, trace and isolation systems in place, countries like Germany have seen increases in reproduction number (*R)* after easing lockdown, with increases to above 1 in June.^3^ South Korea, and China have also recently seen a resurgence in new cases, leading to new localised restrictions being put in place to control the spread of infections.

Easing lockdown when community transmission remains high likely increases the risk of a resurgence of the epidemic but the more precise impacts are insufficiently understood. Several experts, including SAGE, the scientific advisory body to the UK government, cautioned against easing lockdown at this point,^2^ warning that the testing and contact tracing services that are meant to mitigate the impact of easing lockdown, could be overwhelmed and the health service greatly impacted. Nevertheless, the UK has proceeded with easing lockdown with the stated aim of doing so while keeping *R* ≤1. On the 13^th^ May, people who could not work from home were asked to return to work. On the 1^st^ June schools were re-opened, outdoor markets and showrooms opened and households were allowed to meet in socially distanced groups of six. On the 15^th^ June non-essential businesses, including the retail sector, were opened. On the 4^th^ of July, pubs, cafes, and hotels are due to open. However in the week of the 29^th^ June, a surge in cases was reported in Leicester, England, leading to the re-imposition of restrictive measures, and concern that other regions in England may experience similar increases in case numbers.^4^ As of now the government are proceeding with their proposed plan for the 4^th^ July.

Understanding and quantifying the potential impact of lockdown easing measures at this point is crucial to informing public health strategy within England. Here, we model these impacts across a range of plausible scenarios. We use an epidemiological model of COVID-19 spread with Bayesian inference to infer parameters of the epidemic within England using daily death data from the Office of National Statistics (ONS). We estimate the time varying *R* and daily cases, and then use these to forecast cases and deaths in several plausible scenarios in which *R* increases as a result of easing lockdown, particularly focusing on scenarios in which *R* remains ≤1, and contrasting these with elimination strategies that aim to suppress *R* as much as possible.

## Methods

### Data for model development

In order to model the impact of easing lockdown, we need to know the current levels of transmission, and growth parameters of the regional epidemic. Given the limited community testing and case detection in the UK, incident case numbers are likely to be substantially underestimated. We therefore based our model on the number of incident deaths by date of occurrence, which are likely to be more reliable.^5^ Incident deaths are a function of incident cases in the previous weeks and the reproduction rate of the epidemic, and both these parameters can be inferred from the death data.^5^ We included data till the 12^th^ of June for England, as released by the ONS on the 30^th^ of June 2020 (25^th^ week of published data).^6^ These data are based on deaths registered by the 27^th^ of June. As reporting delays mean that more recent deaths are underestimated, we only considered deaths up to the 12^th^ June.

### Primary outcomes

We assessed the excess cumulative predicted cases and deaths, over a 90-day period from the 1^st^ June. We assumed different scenarios of changing *R* at the points of lockdown easing, in comparison with a baseline scenario in which *R* remained constant during this period.

### Estimation of incident cases

Incident cases, and time-varying *R* numbers were estimated using a Bayesian model, similar to that previously described by Flaxman et al,^5^ accounting for the delay between onset of infection and death. The number of infected individuals is modelled using a discrete renewal process, as has been described before.^5^ This is related to the commonly used Susceptible-Infected-Recovered (SIR) model, but is not expressed in differential form.

We modelled cases from 30 days prior to the first day that 10 cumulative deaths were observed in England, similar to previous methods.^5^ The numbers of incident cases for the first 6 days of this period were set as parameters to be estimated by the model (**Supplementary Table 1**). Subsequent incident case numbers would then be a function of these initial cases, and estimated *R* values. We assumed a serial interval (SI) with a lognormal distribution with mean 4.7 and standard deviation (SD) of 2.9 days, as in Nishiura et al ^7^. The SI was discretised as follows:

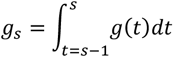

For s=1,2…N, where N is the total number of intervals (each interval being 1 day) estimated. We estimated the distribution for 201 days, to align with the 111 days of data up to the 29^th^ May, plus 90 days of forecasting. Given a SI distribution, the number of infections *c*_*t*_ on a given day *t*, is given by the following discrete convolution function:

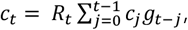

The incident cases on a given day *t*, are therefore a function of *R* at point *t* and incident cases up to time *t-1*, weighted by the distribution of the serial interval.

### Estimation of time-varying reproduction number

The baseline reproduction number (*R*_0_), and the subsequent time varying effective reproduction number (*R*_*t*_) were estimated up to the 12^th^ June. We allowed *R*_*t*_ to change on at least three points: (1) 16^th^ March, when the UK first introduced social distancing measures; (2) 23^rd^ March, when lockdown measures came into place with stay at home instructions and closures of schools and non-essential businesses; and (3) 13^th^ May, the first easing of lockdown. We also considered models in which *R*_*t*_ was allowed to change on the 1^st^ June. Given the limited death data i.e. only up to the 12^th^ June, we were unlikely to be able to estimate changes in *R*_*t*_ after the 13^th^ May with sufficient certainty. Observed deaths from the 1^st^ June are likely to be a function of cases 2-3 weeks prior to this, and were unlikely to reflect changes in *R*_*t*_ from the 1^st^ of June.

### Model selection

We assessed and compared models that allowed *R*_*t*_ to change at the 4 points described above (Model 1), with more flexible models that allowed more frequent changes (Models 2 and 3), as follows:

1. Model 1: 16^th^ March, 23^rd^ March, 13^th^ May and 1^st^ June
2. Model 2: Every week from the beginning of the modelling period, including on the 16^th^ March, 23^rd^ March, 13^th^ May and the 1^st^ June
3. Model 3: 16^th^ March, 23^rd^ March, and 13^th^ May, and every week between the 23^rd^ March and 13^th^ May i.e. during lockdown.

For each model, we used the R package *loo* to calculate expected log pointwise predictive density (ELPD) using Leave-one-out cross-validation (LOO) individually for each left out data point based on the model fit to the other data points. We then calculated between-model differences in ELPDs, to assess whether particular models predicted data better than others, as discussed previously.^8^ As the assumptions in estimation of ELPD may be violated given these are time-series data, and therefore correlated, we also compared the root mean squared errors (RMSE) across models to assess fit. The final model used was arrived upon based on these comparisons.

In addition, we also compared Model 1 (four change points) with models where each of the change points were left out in turn, as done by Dehnig et al,^9^ to assess if these dates do correspond to change points in R_t_.

### Estimation of deaths

Incident deaths from COVID-19 are a function of the infection fatality rate (IFR), the proportion of infections that result in death, and incident cases that have occurred over the past 2-3 weeks. For observed daily deaths (*D*_*t*_) for days *t* ∈ 1, …, n, the expectation of observed daily deaths (*d*_*t*_) is given by:

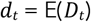

As described in Flaxman et al., we model the number of observed daily deaths *D*_*t*_ as following a negative binomial distribution with mean *d*_*t*_ and variance 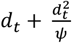, where ψ follows a half normal distribution:

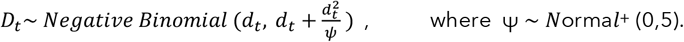

Similar to estimation of incident cases, deaths at time point *t* (*d*_*t*_) were modelled as a function of incident cases up to time *t-1*, weighted by the distribution of time of infection to time of death (*π*). The *π* distribution was modelled as the sum of the distribution of infection onset to symptom onset (the incubation period), and the distribution of symptom onset to death. As has been previously done,^5^ both of these were modelled as gamma distributions with means of 5.1 days (coefficient of variation 0.86) and 18.8 days (coefficient of variation 0.45), respectively as follows:

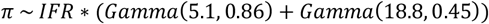

IFR was assumed to be 1.1%, based on the most recent estimates from the University of Cambridge MRC Nowcasting and Forecasting model.^10^

To discretise this distribution, we estimated the probability of death within each discrete time interval (1 day), conditional on surviving previous intervals. First we calculate the hazard (*h*_*t*_) the instantaneous probability of failure (i.e. dying) within a time interval, as follows:

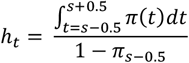

As the denominator excludes individuals who have died, this ensures that *h*_*t*_ is calculated only among those surviving. The probability of survival within each interval is:

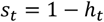

The cumulative survival probability of surviving up to the interval *t-1* is therefore:

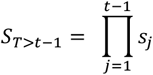

, where *T* is the time of death of an individual. In other words the cumulative probability of survival up to interval *t* is simply the product of survival within each interval up to *t-1*, where the probability of survival within each interval (*s*_*t*_) is *1-h*_*t*_, where *h*_*t*_ is the probability of dying within that interval.

Given this, we now estimate the probability of death within interval *t*, conditional on surviving up to *t-1* as:

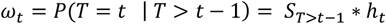

Here *ω* represents the discretised distribution of infection onset to death, with the probability of death within interval *t* conditional on surviving previous intervals. Deaths can therefore be calculated as a function of incident cases of infection within previous intervals, as follows:

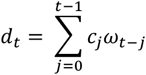

Here, the number of deaths within interval *t* (on a given day) is a sum of the number of daily cases up to the previous day, with previous cases weighted by the discretised probability distribution of time from onset of infection to death.

### Estimated parameters and model priors

We estimated the set of model parameters θ={c_*1-6*_, *R*_*0*_, *R*_*t*_, ϕ, ***τ***} using Bayesian inference with Markov-chain Monte-Carlo (MCMC) (**Supplementary Table 1**). We estimated the number of cases in the first six days of the modelled period, as subsequent cases are simply a function of cases on these days, the SI, and *R*_*t*_. As described above, *R*_*0*_ was constrained up to the 16^th^ March and then again after the 13^th^ of May. For the period prior to 16^th^ March, we assigned a normal prior for *R*_*0*_ with mean 2.5 and SD 0.5. For the period that *R*_*t*_ was allowed to vary i.e. every week from the 16^th^ of March till the 13^th^ of May, we assigned a normal prior with a mean 0.8 and SD 0.25. These priors are based on estimates of time changing *R*_*t*_ from the University of Cambridge MRC biostatistics nowcasting and forecasting models^10^ and SAGE estimates of *R*,^11^ and consistent with Flaxman et al.^5^ For the number of cases on day 1, we assigned a prior exponential distribution:

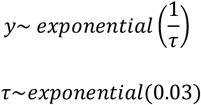

where

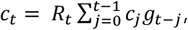

### Model estimation

Parameters were estimated using the Stan package in R with Markov chain Monte Carlo (MCMC) algorithms used to approximate a posterior distribution of parameters by randomly sampling the parameter space. We used 4 chains with 1000 warm up samples (which were discarded), and 3000 subsequent samples in each chain (12,000 samples in total) to approximate a posterior distribution using the Gibbs Sampling algorithm. From these we obtained the best-fit values and the 95% credible intervals for all parameters. We used these parameters to estimate the number of incident cases and deaths in England. We examined the fit of the model predicted deaths to the observed daily deaths from the ONS, and also the consistency of the model parameters with known values in the literature, estimated from global data. We assessed the distribution of *R-hat* values for all parameters, to assess convergence between chains.

### Sensitivity analyses

We carried out sensitivity analyses using broader, and uninformative priors for *R*_*0*_ and *R*_*t*_, to examine the sensitivity of *R*_*t*_ estimates to prior specification. We also examined the impact of the SI by comparing the baseline model (SI of mean 4.7 and SD 2.9 days), with a longer SI modelled as a gamma distribution with mean 6.5 and coefficient of variation of 0.72, as estimated by Chan et al.^12^

### Forecasting cases and deaths

All forecasts were carried out up to 90 days (29^th^ August 2020) after the 1^st^ of June. We considered a set of scenarios in which *R*_*t*_ increased from baseline on the 1^st^ of June and then remained constant, as well as those in which further increases in *R*_*t*_ occur on the 15^th^ June and the 4^th^ July (**Figures 3a, 4a** and **5a**). We considered an increase in *R*_*t*_ of up to 0.25 in increments of 0.05, this being a plausible degree of change in response to easing lockdown, based on the empirical data from other countries,^3,13^ as well as the modelling by UK SAGE.^14^ Finally, for comparison with a strategy of elimination, namely suppressing *R*_*t*_ to the lowest level possible before easing lockdown measures, as has been done South Korea, New Zealand and Australia, we also modelled scenarios with *R*_*t*_ values of 0.6 and 0.7.

**Figure 1:**
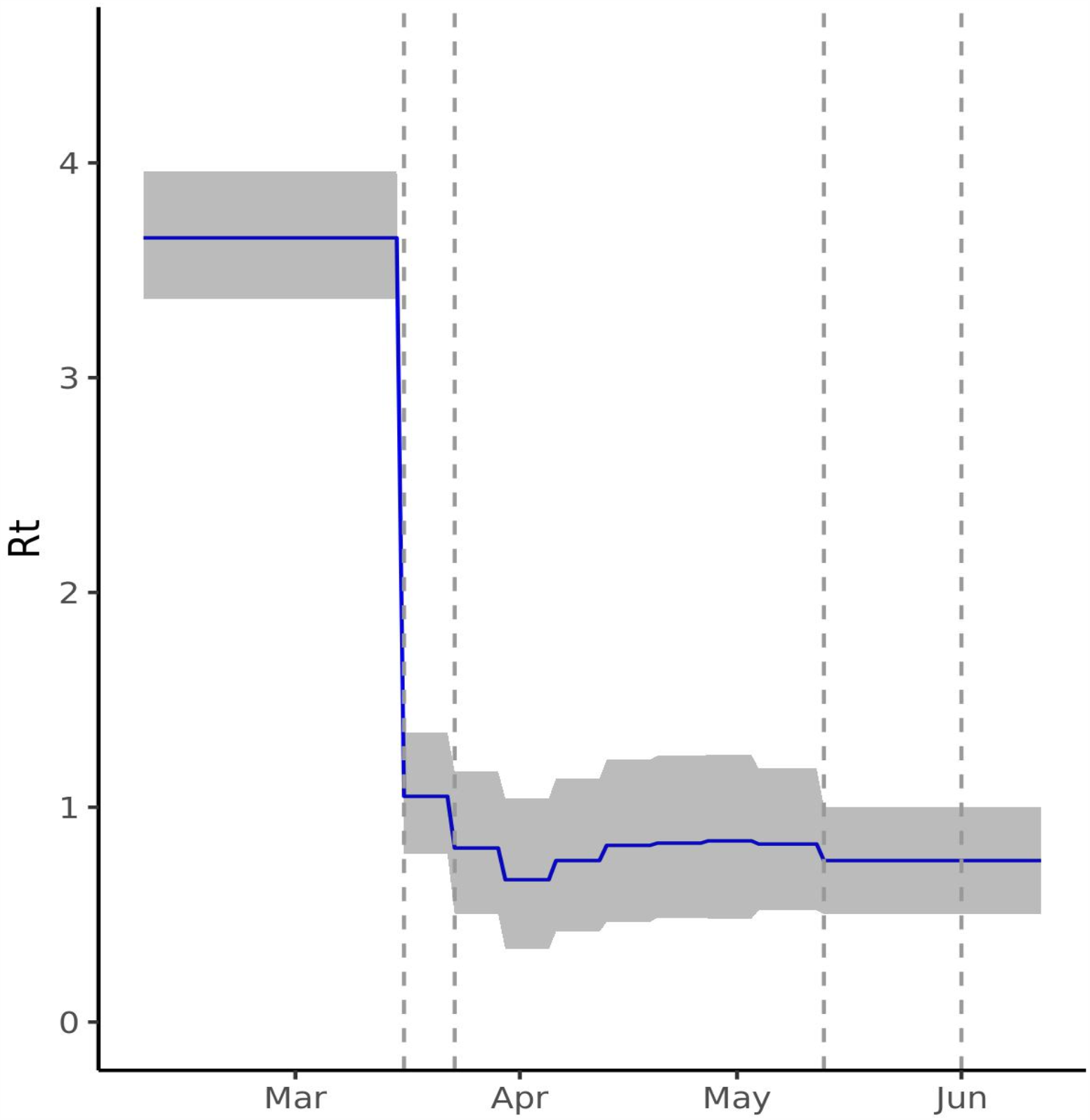
Estimated time-varying reproduction number (*R*_*t*_) for England. The figure shows the *R*_*t*_ estimated by Model 3 (blue) with 95% credible intervals (grey) with a serial interval of mean 4.7 and SD 2.9 days. From 3.65 (CI 3.36-3.96), *R*_*t*_ drops on the 16^th^ March and 23^rd^ March (indicated by vertical dashed lines) when social distancing and lockdown were instituted, reaching a low of 0.66 (95% CI 0.34-1.04) in the week of the 30^th^ March. The last estimated *R*_*t*_ is 0.75 (95% CI 0.50-1.00) following the 13^th^ May.

**Figure 2:**
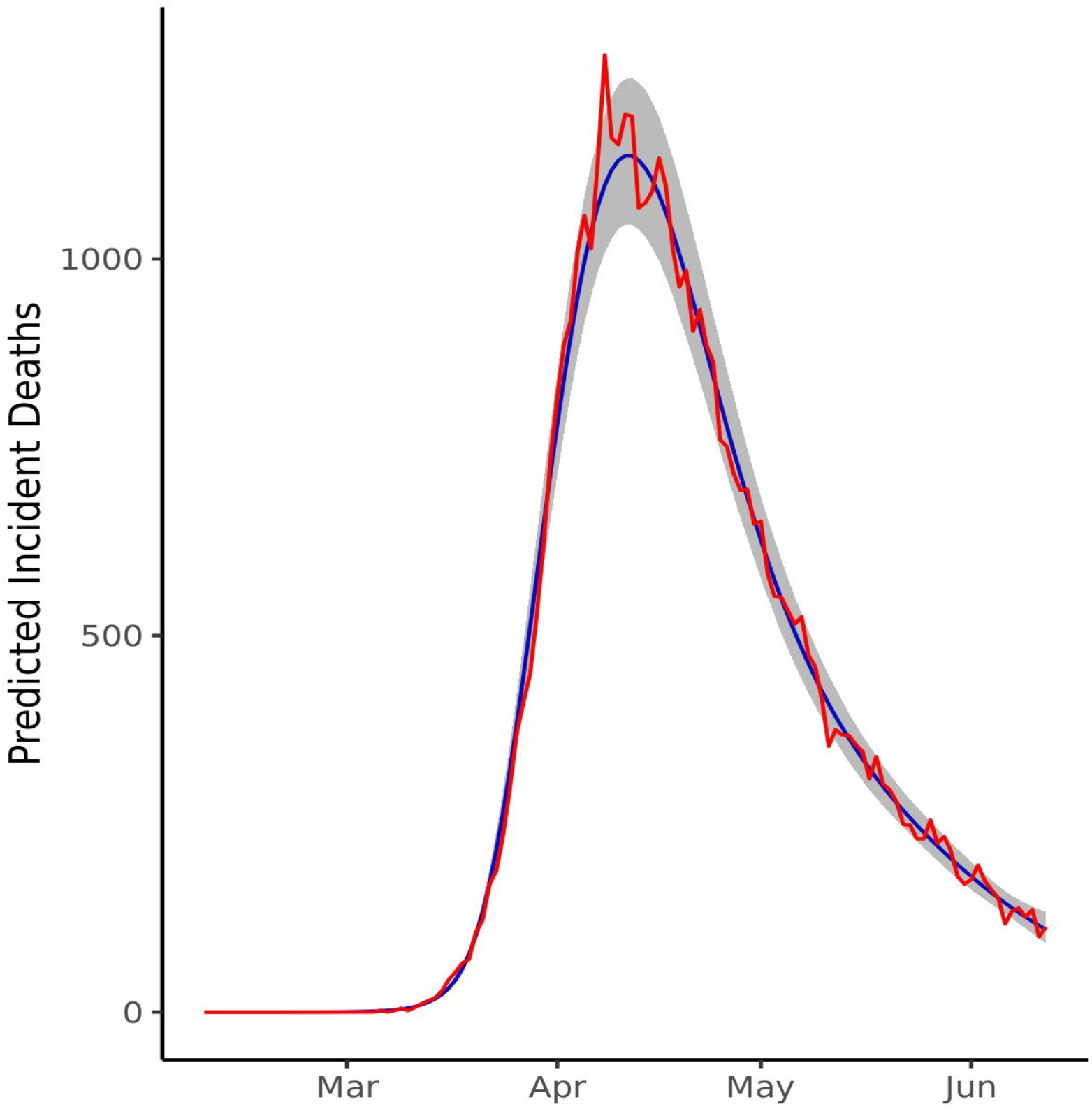
Model fit to observed death data. Daily deaths predicted by Model 3 (blue) with 95% credible intervals (grey) show a good fit to the observed deaths from the ONS (red)

**Figure 3.**
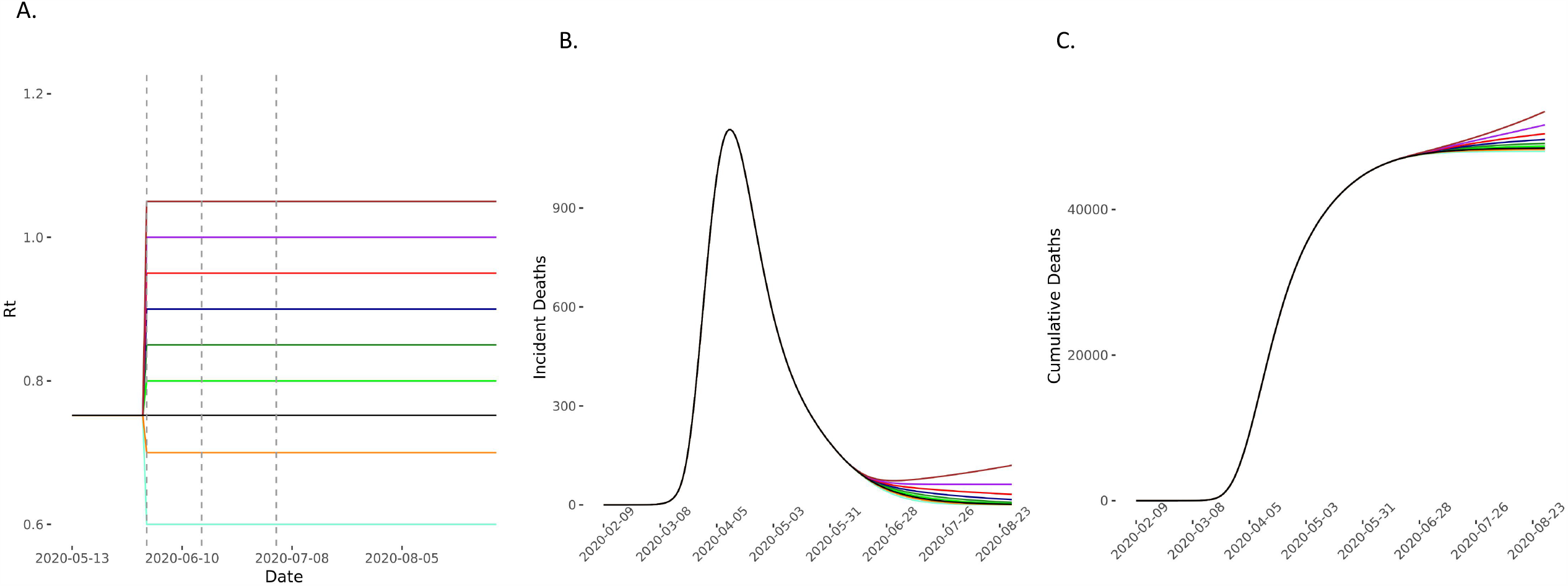
Predicted deaths with *R*_*t*_ increasing on 1^st^ June. (A) The model compared scenarios in which *R*_*t*_ increases to 0.80 (light green), 0.85 (green), 0.90 (dark blue), 0.95 (red), 1 (purple) and 1.05 (brown) and then remains constant for the 90-day forecasting period. The comparator baseline scenario is of *R*_*t*_ remaining at 0.75 (black) and two elimination strategies of *R*_*t*_ reducing to 0.7 (yellow) and 0.6 (light blue) were also considered. Vertical dashed lines represent time-points of easing lockdown. (B), (C) the incident and cumulative deaths increase in all scenarios in which *R*_*t*_ increases and reduces in the two elimination scenarios.

**Figure 4.**
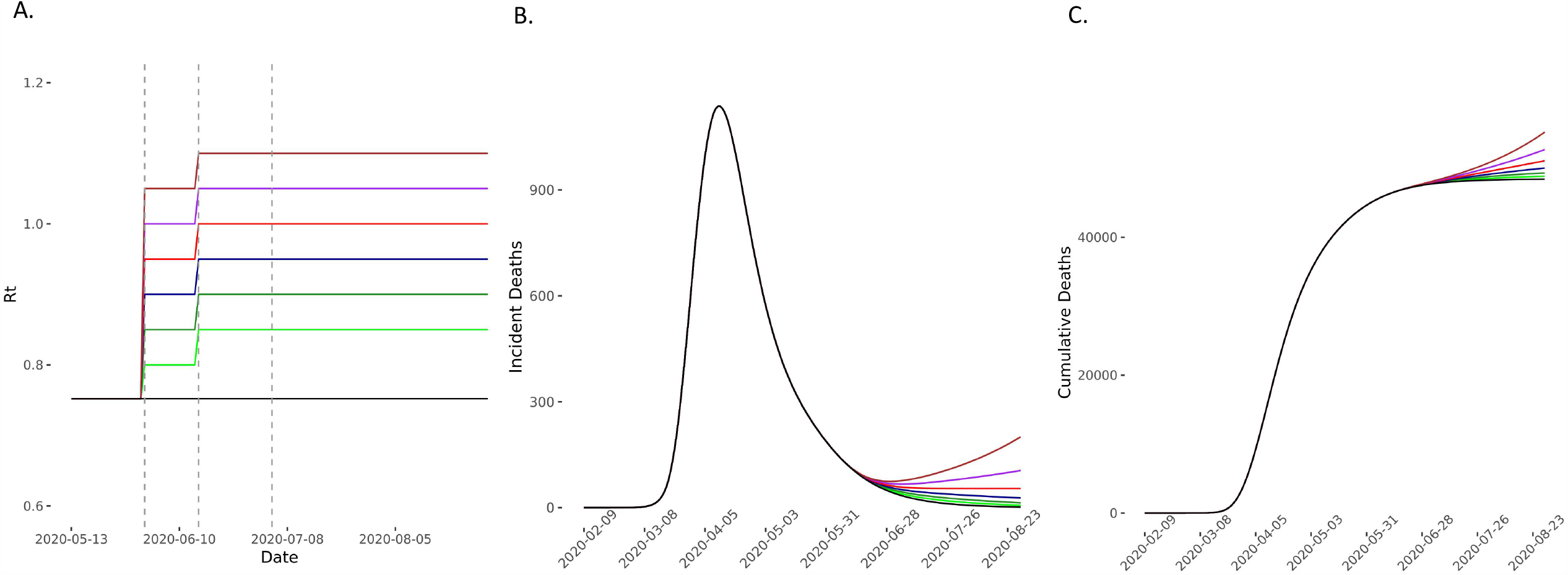
Predicted deaths in scenarios of *R*_*t*_ increase on 1^st^ and 15^th^ June compared with baseline scenario. (A) The model compared scenarios in which *R*_*t*_ increases to 0.80 (light green), 0.85 (green), 0.90 (blue), 0.95 (red), 1 (purple) and 1.05(brown) and then further by 0.05 on the 15^th^ June and then remaining constant for the 90-day forecasting period. The comparator baseline scenario is of *R*_*t*_ remaining at 0.75 (black). Vertical dashed lines represent time-points of easing lockdown. (B), (C) The incident and cumulative deaths increase in all scenarios in which *R*_*t*_ increases.

**Figure 5.**
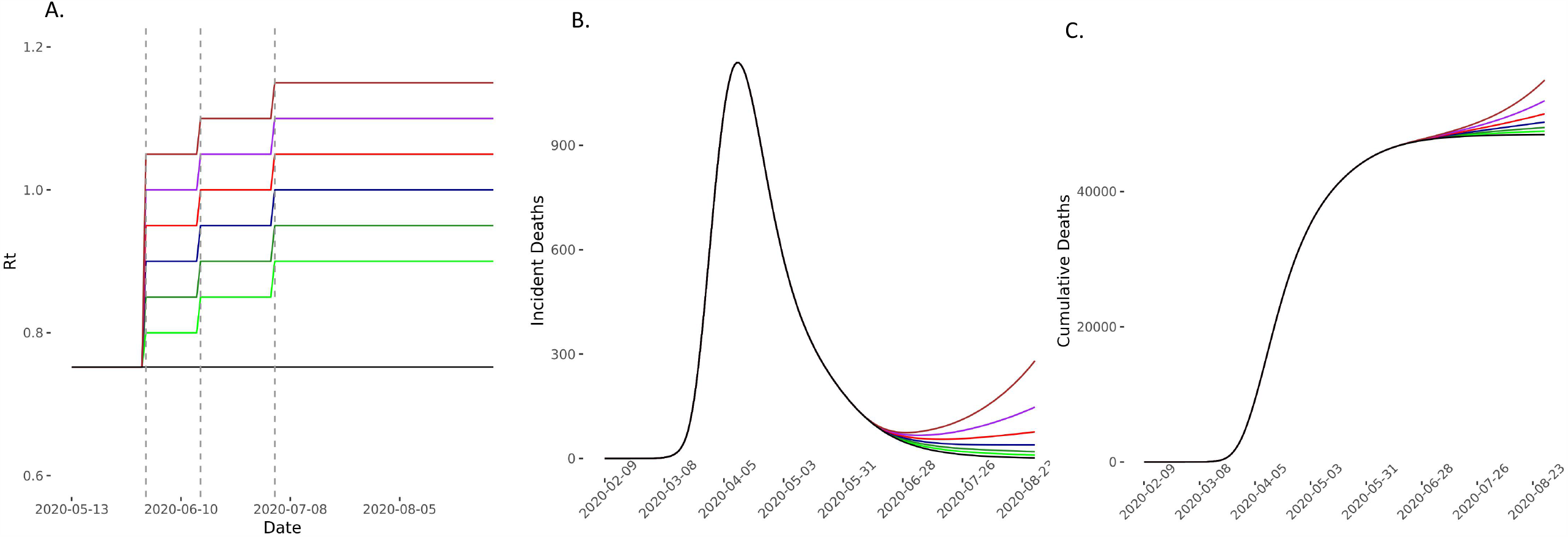
Predicted deaths in scenarios of *R*_*t*_ increase on 1^st^ June, 15^th^ June and 4^th^ July compared with baseline scenario. (A) The model compared scenarios in which *R*_*t*_ increases to 0.80 (light green), 0.85 (green), 0.90 (blue), 0.95 (red), 1 (purple) and 1.05(brown) and then further by 0.05 on the 15^th^ June and then again by 0.05 on the 3^rd^ July before remaining constant for the 90-day forecasting period. The comparator baseline scenario is of *R*_*t*_ remaining at 0.752 (black). Vertical dashed lines represent time-points of easing lockdown. (B), (C) The incident and cumulative deaths increase in all scenarios in which *R*_*t*_ increases.

For each of these scenarios, we predicted the number of incident cases, and incident deaths, using the functions from the inference model above. Briefly cases are a function of *R*_*t*_, incident cases on previous days and the SI discretised distribution:

Deaths are a function of incident cases over previous weeks, and the distribution of onset of infection to death times:

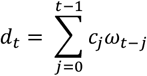

All scenarios were compared to a baseline scenario of no change in *R*_*t*_ from the 13^th^ of May onwards.

## Results

### Model selection and model inferences

Model 3, which allowed weekly changes in *R*_*t*_ during lockdown, produced the best fit to the data (**Supplementary Table 2**), with estimation of fewer parameters compared with Model 2. This was therefore used as the primary model and unless otherwise stated, all inferences described subsequently are from this model.

We infer *R*_*0*_ of 3.65 (95% credible intervals (CI) 3.36-3.96), consistent with previous estimates within the UK.^5^ The *R*_*t*_ is estimated to have declined substantially following initiation of social distancing, and lockdown measures, reaching a low of 0.66 (95% CI 0.34-1.04) during the week 30^th^ March-5^th^ April 2020. The most recent *R*_*t*_ from the 13^th^ of May is estimated as 0.752 (95% CI 0.50-1.00) (**Figure 1**). The alternative models allowing change of *R*_*t*_ on the 1^st^ of June inferred a very similar *R*_*t*_ for the 1^st^ -12th June suggesting that there was insufficient data to accurately infer any changes to *R*_*t*_ following the easing of lockdown on 1^st^ June. On examining the impact of constraining *R*_*t*_ on model fit at any of the 4 change points, this appears greatest for the 16^th^ March (when social distancing measures were put into place) (**Supplementary Table 3**) with only modest impacts on model fit of constraining *R*_*t*_ on 23^rd^ March and 13^th^ May, and no impact on constraining *R*_*t*_ on the 1^st^ June.

The model showed a good fit to the observed distribution of deaths up to the 12^th^ June (**Figure 2**). *Rhat* estimates were < 1.05 for all estimated parameters (**Supplementary Figure 1**). Leave one out cross-validation also supported a good model fit, with the shape parameter k<0.5 for all values (**Supplementary Figure 2**). The median number of incident cases inferred on the 1^st^ June was 4,317/day (95% CI 2,062-8,155), which is broadly consistent with the estimates from the ONS survey for England based on a random sample of the population within the same time period.

### Forecasts of lockdown easing scenarios

In the baseline forecasting scenario where *R*_*t*_ remains constant (Rt_est_=0.75) through the 90-day forecasting period (1^st^ June to 29^th^ August 2020), the model predicts 48,501 (46,170-50,989) cumulative deaths in England (**Supplementary Table 4**). By comparison, the ONS reported 46,539 cumulative deaths up to 12^th^ June in England (registered up to 27^th^ June).

In the scenarios where *R*_*t*_ increases on the 1^st^ of June and then remains constant, for increases from the median 0.75 to 0.80, 0.85, 0.90, 0.95 and 1, the model predicts median excess deaths of 257 (95% CI 108-492), 632 (95% CI 265-1,208), 1,173 (95% 493-2,240), 1,971(95% 828-3,764) and 3,174 (95% CI 1,334-6,060) respectively. Increases of *R*_*t*_ to 1.05 and 1.1, with resultant exponential growth, lead to excess median deaths of 5017 (95% CI 2,109-9,578), and 7,878 (3,313-15,037) respectively (**Figure 3** and **Supplementary Table 4**).

In scenarios where *R*_*t*_ increases on the 1^st^ June, 15^th^ June and 4^th^ July, we find that compared to the baseline scenario, modest increases of *R*_*t*_ to 0.80, 0.85, 0.90, on these dates respectively would lead to 508 (95% CI 213-972) excess deaths. If *R*_*t*_ increases to 0.90, 0.95 and 1 at these time points, then excess estimated deaths increase to 1,848 (95% CI 776-3,534). In these scenarios *R*_*t*_ remains ≤1 (**Figures 3-5** and **Supplementary Table 4**). Increases of *R*_*t*_ above 1 at any point of results in rapid increases in cases, and deaths, predicting a second wave of the epidemic within England (**Figure 4-5** and **Supplementary Table 4**).

Even in a conservative scenario where *R*_*t*_ increases from 0.75 to 0.80 on the 1^st^ of June and then remains constant thereafter, the model predicts an excess of 26,447 (95% CI 11,105-50,549) cumulative cases over 90 days. On the other hand, the scenario with the largest changes in *R*_*t*_, but still remaining ≤1, predicts an excess of up to 421,310 (95% CI 177,012-804,811) (**Figures 6-8** and **Supplementary Table 4**).

**Figure 6.**
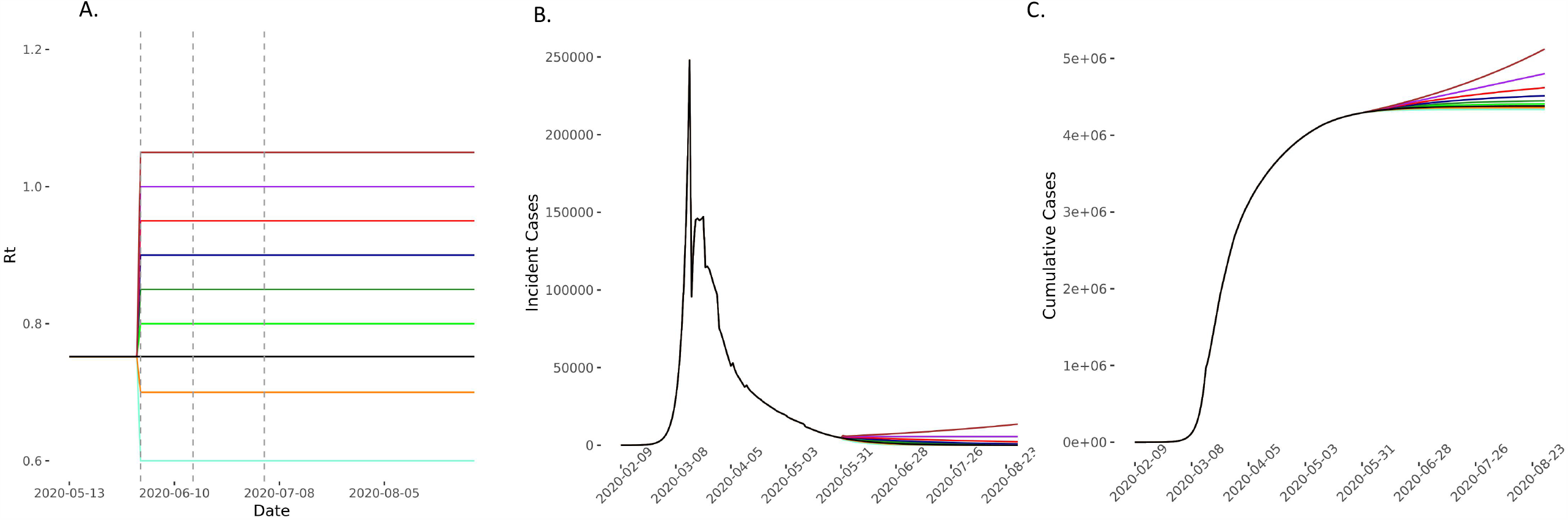
Predicted cases in scenarios of *R*_*t*_ increase on 1^st^ June compared with baseline and elimination scenarios. (A) The model compared scenarios in which *R*_*t*_ increases to 0.80 (light green), 0.85 (green), 0.90 (dark blue), 0.95 (red), 1 (purple) and 1.05(brown) and then remains constant for the 90-day forecasting period. The comparator baseline scenario is of *R*_*t*_ remaining at 0.752 (black) and two elimination strategies of *R*_*t*_ reducing to 0.7 (yellow) and 0.6(light blue) were also considered. Vertical dashed lines represent time-points of easing lockdown. (B), (C) the incident and cumulative cases increase in all scenarios in which *R*_*t*_ increases and reduces in the two elimination scenarios.

**Figure 7.**
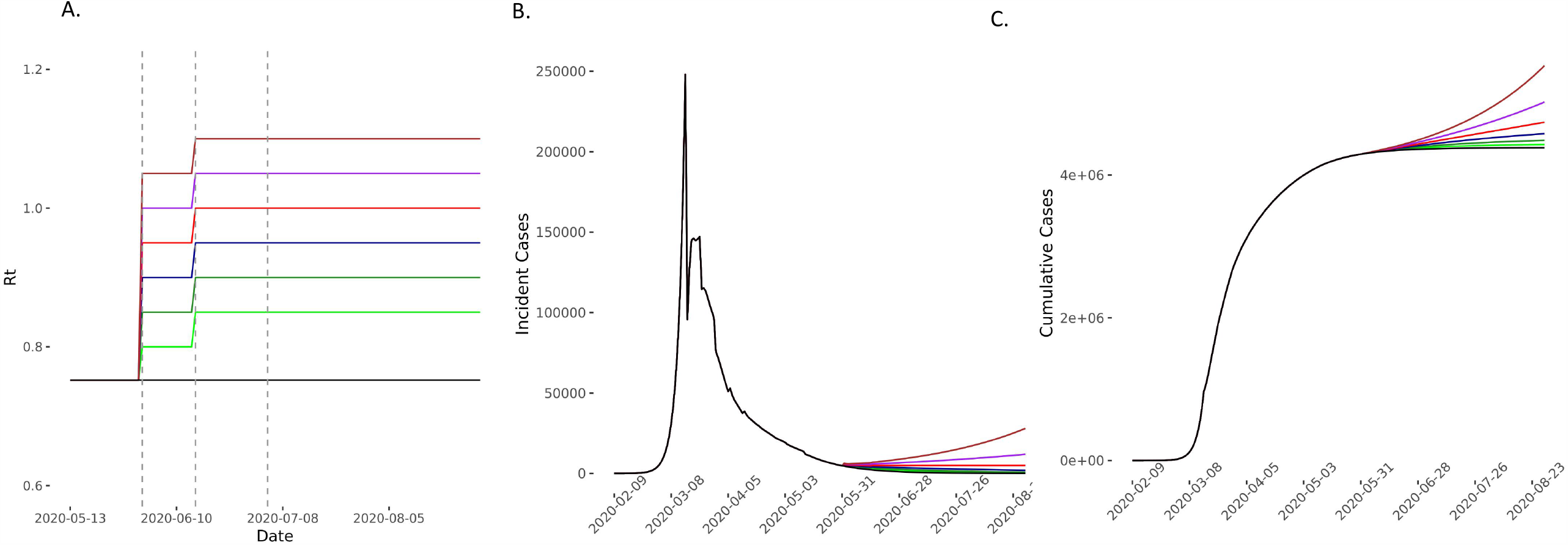
Predicted cases in scenarios of *R*_*t*_ increase on 1^st^ June and 15^th^ June compared with the baseline scenario. (A) The model compared scenarios in which *R*_*t*_ increases to 0.80 (light green), 0.85 (green), 0.90 (blue), 0.95 (red), 1 (purple) and 1.05(brown) and then further by 0.05 on the 15^th^ June and then remaining constant for the 90-day forecasting period. The comparator baseline scenario is of *R*_*t*_ remaining at 0.752 (black). Vertical dashed lines represent time-points of easing lockdown. (B), (C) The incident and cases increase in all scenarios in which *R*_*t*_ increases.

**Figure 8.**
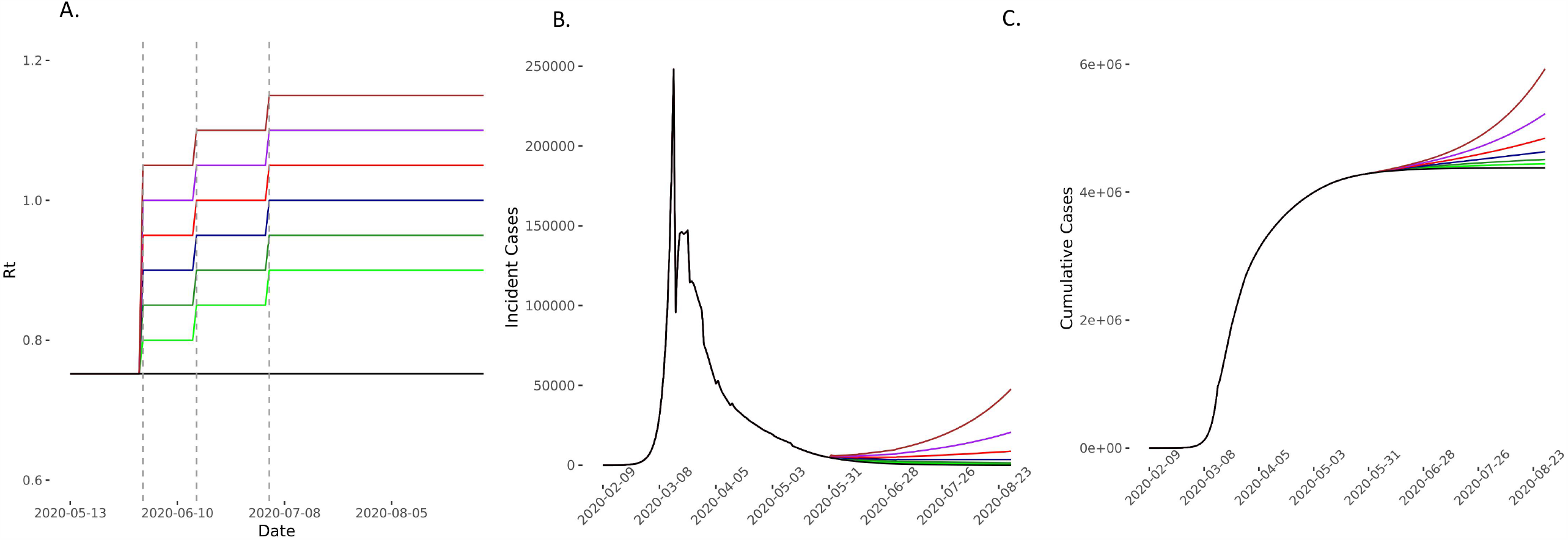
Predicted cases in scenarios of *R*_*t*_ increase on 1^st^ June and 15^th^ June and 4^th^ July compared with the baseline scenario. (A) The model compared scenarios in which *R*_*t*_ increases to 0.80 (light green), 0.85 (green), 0.90 (blue), 0.95 (red), 1 (purple) and 1.05(brown) and then further by 0.05 on the 15^th^ June and then again by 0.05 on the 3^rd^ July before remaining constant for the 90-day forecasting period. The comparator baseline scenario is of *R*_*t*_ remaining at 0.752 (black). Vertical dashed lines represent time-points of easing lockdown. (B), (C) The incident and cumulative cases increase in all scenarios in which *R*_*t*_ increases.

### Forecasts from an elimination scenario

Compared to the baseline scenario of *R*_*t*_ staying at 0.75, we find that maintaining *R*_*t*_ at 0.60 and 0.70 would result in 44,302(95% CI 84684-18600) and 19,968 (95% CI 38168-8384) fewer cumulative cases, and 462 (95% CI 194-884) and 204 (95% CI 389-86) fewer deaths over the modelled 90-day period, respectively **(Figure 3, Figure 6, Supplementary Table 4**).

### Robustness of model in sensitivity analyses

Using uninformative (no prior specified) priors for *R*_*t*_ did not materially alter the median estimates of *R*_*t*_, although uncertainty around estimates was predictably increased (**Supplementary Figure 3**). This suggests our estimates are robust to the priors specified.

Using a longer SI leads to an increase in the estimated *R*_*0*_, although subsequent estimates following easing of lockdown remain broadly similar (**Supplementary Figure 4**). This model is comparable to the primary model with regard to fit to observed deaths (**Supplementary Figure 5**) and in predicted excess deaths and cases in scenarios where *R*_*t*_ increases (**Supplementary Table 5**).

## Discussion

In this paper we describe a Bayesian model for inferring incident cases and reproduction numbers from daily death data, and for forecasting the impact of future changes in *R*. Our findings provide important quantification of the likely impact of relaxing lockdown measures in England, and to our knowledge, this is the first study to comprehensively assess this through several plausible scenarios. We show that even in scenarios in which *R* remains ≤1 (in line with the UK government’s stated aim), small increases in *R*_*t*_ from lifting lockdown measures, can lead to a substantial excess of deaths with 3,174 (95% CI 1,334-6,060) in the most severe scenario modelled.

Our model inferences are robust to modelling assumptions of serial interval distribution, and specified priors. Our estimated *R*_*t*_ of 0.75 following 13^th^ May is consistent with estimates from the SAGE group advising government.^11^ We have assessed increases in *R*_*t*_ that are entirely plausible, given the data from other European countries that have started easing lockdown.^3^ Our model predicts a substantial excess of cases and deaths in scenarios where *R* remains ≤1. Rises in *R*_*t*_ above 1 would lead to exponential increases in cases, and subsequently deaths. In contrast, we show that pursuing an elimination strategy where *R*_*t*_ would be suppressed to 0.6 or 0.7 could prevent a median estimated 462 and 204 deaths, and 44,302, and 19,968 cases, respectively.

Unlike other European countries, the UK began to ease lockdown when community transmission was still high with an estimated incidence of infection of >8000 cases and >300 deaths being observed per day in England. In Denmark and Germany some of the increases in *R* since easing lockdown, have likely been mitigated by the low levels of transmission at the point of easing lockdown. Another important factor may be the use of aggressive case detection and contact tracing approaches, which the UK seems unlikely to have fully operational till later this year, and the existing system is at risk of being overwhelmed by major increases in incident cases. Given the lack of comprehensive testing, the UK’s current estimates of *R*_*t*_ rely on incident deaths (as used by the MRC Nowcasting and Forecasting model)^10^, which means that changes in *R*_*t*_ reflect changes in community transmission from a median of 2-3 weeks ago.^11^ With lockdown being eased in 2-weekly steps, this means that by the time we detected the impact of one step, the next one would already have been instituted so mitigating these impacts would be extremely challenging. The UK SAGE has also expressed concerns that increases in *R* up to 1.2 may continue undetected for longer periods of time.^14^ These concerns have been borne out by the recent surge in cases observed in Leicester,^4^ where the increase in case numbers were only detected two weeks after the event. Our findings strongly suggest that despite small increases in *R*, we would likely see substantial increases in cases and deaths, which may be detected too late to mitigate impact of the lockdown easing measures that led to these. This is particularly important when we consider the impact on health services, which have managed to deal with the pandemic by suspending much of routine healthcare, which is likely to substantially increase indirect causes of deaths from cancer, and cardiovascular disease. This also has important implications for population health, as we observe multi-system long-term sequelae among those infected with COVID-19.

We acknowledge some important limitations of our model. The first is that it is based on a back calculation of cases based on incident deaths, which are likely to underestimated due to reporting delays and underreporting. Second, our model is reliant on inferring cases, and reproduction numbers, which depend on the assumed distributions of the serial interval, and the time of onset to death distributions. While we have based our assumptions on the literature, misspecification of these would influence our estimates. While we have evaluated this, greater deviations from true estimates would make our forecasting less reliable. Third, similar to Flaxman et al, our model uses the IFR as a multiplier for the distribution of time from infection to death, in the absence of reliable population level case fatality rates (CFR). While this would not affect the estimation of deaths, if the CFR were higher (due to large proportions of cases being asymptomatic), then the predicted case numbers would be overestimated by our model. We note, however that the estimate of IFR we used (1.1%) is consistent with the CFR estimated in previously from Beijing.^15^ We have also, for simplicity, assumed that IFR remains constant throughout the pandemic and the forecasting period, and this may not reflect complex heterogeneity in IFR over time. Finally, we do not consider the impact of mitigatory measures in our current modelling. However mitigatory measures are likely to be implemented with significant delays from when community transmission increases, namely when changes in *R* are detected. If such measures, like re-introducing lockdown, or school closures, were re-implemented, they may reduce the impact of the modelled scenarios.

In summary, we show that increases in *R*_*t*_ as a result of easing lockdown would have a substantial impact on incident transmission and deaths for even modest increases that still maintain *R*_*t*_ ≤ 1. We argue for a more cautious approach with a focus on elimination, by reducing *R*_*t*_ and incident cases to low levels prior to easing lockdown measures and then too with careful monitoring.

## Data Availability

All data on daily deaths used in this study were taken from the Office of National Statistics website (https://www.ons.gov.uk/peoplepopulationandcommunity/birthsdeathsandmarriages/deaths/datasets/weeklyprovisionalfiguresondeathsregisteredinenglandandwales). The code for the model is available at: https://github.com/dgurdasani1/lockdownsim

## Contributors

DG conceived the study and designed the model with NS. DG programmed the model and made the figures. HZ and NS consulted on the model design. All authors interpreted the results, contributed to writing the Article, and approved the final version for submission.

## Data sharing

All data on daily deaths used in this study were taken from the Office of National Statistics website (https://www.ons.gov.uk/peoplepopulationandcommunity/birthsdeathsandmarriages/deaths/datasets/weeklyprovisionalfiguresondeathsregisteredinenglandandwales).

The code for the model is available at: https://github.com/dgurdasani1/lockdownsim

## Acknowledgements

HZ is partly funded by the Bernard Wolfe Health Neuroscience Fund at the University of Cambridge. NS is funded by a Strategic Award from the Wellcome Trust. DG is funded by the UKRI/Rutherford HDR-UK fellowship programme (reference MR/S003711/2).

We would like to particularly acknowledge and thank Flaxman et al and the team at Imperial College London for making their code available for us to use.

